# DRUM-PD/HD: The Impact of a Pilot Group Drum-Based Music Therapy Intervention on Quality of Life and Motor Symptoms in Parkinson Disease and Huntington Disease

**DOI:** 10.1101/2023.05.04.23289514

**Authors:** Lavinia Wainwright, Kyurim Kang, Gabriel Dayanim, Chris Bedell, Kerry Devlin, Diane Lanham, Gerson Suarez-Cedeno, Jason Armstrong Baker, Serap Bastepe-Gray, Jee Bang, Alexander Pantelyat

## Abstract

Rhythm-based therapeutic interventions have widely been used in patients with neurologic disorders to address motor and quality of life outcomes. Although group drumming has been explored in several pilot studies of patients with Parkinson disease (PD) and Huntington disease (HD), caregiver burden and their quality-of-life outcomes have received less attention. Therefore, we aimed to evaluate the impact of group drumming on quality of life and motor symptoms in PD and HD patients as well as their caregivers. A total of 17 participants (PD = 6, caregivers of PD = 6, HD = 3, caregivers of HD = 2) attended the 60-minute drum-based group music therapy intervention twice a week for 12 weeks. Participants were assigned to two drumming intervention groups based on their diagnosis:1) patients diagnosed with PD and their study partners, 2) patients diagnosed with HD and their study partners. In group drumming sessions, both patients and caregiver participants utilized a variety of percussion instruments based on their personal preferences or physical abilities to facilitate movement skills and group cohesion. They were asked to complete questionnaires about quality of life and motor functioning at baseline, 6 weeks, 12 weeks, and 18 weeks (6 weeks post-intervention completion). Caregivers burden scores were also collected at these time points. Furthermore, all participants completed simple exit interview questionnaires in their follow-up visit. The PD participants and their caregivers showed an opposite trend in social role satisfaction. From baseline to 6 weeks, there were significant differences in social role satisfaction between PD participants and PD caregivers where PD participants reported a decrease in social satisfaction (*Mean* = -2.30, *Standard Deviation* = 1.64) while PD caregivers experienced an increase (*M* = 3.80, *SD* = 3.08), *p* = .02. In contrast, a different pattern was shown from 12 weeks to 18 weeks, where PD participants showed an increase in social satisfaction (*M* = 2.53, *SD* = 1.29), while PD caregivers demonstrated a decrease in social satisfaction (*M* = -2.10, *SD* = 3.35), *p* =.03. Drumming in a group setting may serve as an effective tool to enhance movement and promote social cohesion through rhythmic auditory-motor entrainment, thereby supporting quality of life in PD; further studies in HD are indicated as well.

## Background

Studies indicate that rhythmic entrainment (the process in which the brain and body temporally synchronize with external rhythmic auditory cues) to an auditory stimulus such as live music or a metronome may create functional changes in the timing, spatial and force parameters of movements in patients with movement disorders by activating structures within key motor networks in the brain (Braunlich et al., 2019; Grahn & Watson, 2013; Koshimori & Thaut, 2018; Nombela et al., 2013; Thaut, 2015). Rhythm in particular may provide a foundational structure for the damaged brain in patients with movement disorders by priming the motor system and increasing subsequent motor response quality (Thaut et al., 1996, 2015).

Rhythm-based music therapy interventions have widely been used in patients with neurologic disorders to address motor and quality of life outcomes (Daveson, 2007; Davis & Magee, 2001; Magee et al., 2017; Yoo & Kim, 2016). The impact of music therapy interventions on caregiver-patient interactions for patients diagnosed with dementia and neurologic disorders, as well as those receiving end-of-life care, has also been explored for several decades (Brotons & Marti, 2003; Clair & Ebberts, 1997; Hanser et al., 2011; Hilliard, 2006).

Psychological distress may often contribute decreased quality of life in patients with Parkinson Disease (PD), and depression may worsen PD motor symptoms (Chapuis et al., 2005; Gomez-Esteban et al., 2007; Helder et al., 2001; Schrag et al., 2000). Rhythm-based group music therapy programs or music-based interventions may provide opportunities for patients to address motor and quality of life needs in a social setting with peers experiencing similar challenges. Research also indicates that group music therapy interventions combined with standard care are feasible for patients with depression and may be associated with improvements in mood and alteration of stress-related hormones (Castillo-Pérez et al., 2010; Erkkilä et al., 2011; Maratos et al., 2008).

Previously, the Drum-PD pilot study found that compared to patients with PD who pursued usual care (n=10), those who attended twice-weekly West African drumming classes for 6 weeks (n=8) had significantly improved self-reported total Parkinson’s Disease Questionnaire-39 quality of life scores and, from baseline to 12 weeks, demonstrated trend-level improvements in walking (Pantelyat et al., 2016). In Huntington disease, Metzler-

Baddeley et al., (2014) explored drumming and rhythm exercises targeting early dysexecutive problems, such as difficulties in sequence and reversal learning, response speed, timing, and dual tasking in nine patients with early HD and one presymptomatic individual. Five patients completed two months of training and improvements in executive function and magnetic resonance imaging (MRI) changes in white matter microstructure, notably in the genu of the corpus callosum (which connects prefrontal cortices of both hemispheres), were observed. Casella et al., (2020) assessed the impact of two months of the same drumming protocol as in the above study in 8 patients with HD and 9 controls. They found improvements in drumming performance in both groups and also detected a significantly greater change in a MRI measure of myelin integrity (macromolecular proton fraction from quantitative magnetization transfer imaging sequences) for HD patients relative to controls post-training; these changes were found in two of three assessed portions of the corpus callosum and in the white matter pathway connecting the right supplementary motor area and putamen. However, though patients improved their drumming and executive function, these improvements did not correlate with microstructural MRI changes.

It should be noted that the above studies did not assess caregiver burden, which is substantial in care partners of patients with both PD and HD (Hergert & Cimino, 2021; Mosley et al., 2017; Pickett Jr et al., 2007; Yu et al., 2019). In addition, no studies known to the authors involved patients and their caregivers side by side in the same drumming intervention.

The purpose of this pilot study was to investigate whether a drum-based music therapy protocol called *The Armstrong Rhythm Method* ^***SM***^ would impact functional motor outcomes for patients diagnosed with PD or HD, as well as quality of life for both these patients and their caregivers.

## Methodology

### Participants

Participants with a clinical diagnosis of PD (*n*=6) according to MDS criteria (Postuma et al., 2015); a genetic diagnosis of HD (*n*=3); and their caregivers (*n*=8; one caregiver was shared by 2 related HD participants) were enrolled in this study. Eligibility criteria included the absence of significant injury or co-morbid diagnosis affecting the upper extremities for instrument play (e.g., broken bones, sprains, severe arthritis, conditions involving paresis), no active psychosis and no other neurological conditions. Also, only participants who scored better than 17 out of 30 on the Montreal Cognitive Assessment (MoCA) (Nasreddine et al., 2005) were enrolled; patients with notably impaired cognitive performance were thought to be unlikely to experience optimal benefit from study interventions due to difficulty following the therapist’s instructions.

For PD, severity was assessed with the MDS-Unified Parkinson Disease Rating Scale (MDS-UPDRS) (Goetz et al., 2008) and Hoehn & Yahr staging. Disease severity for HD was assessed with the Unified Huntington Disease Rating Scale (UHDRS)-Motor and Unified HD Rating Scale-Total Functional Capacity (Kieburtz et al., 1996).

In accordance with the declaration of Helsinki, the study coordinator explained the intervention protocol to all participants and written informed consent was obtained prior to participation in the study. The study protocol was approved by the Institutional Review Board at the Johns Hopkins Medical Institutions (Baltimore, MD; IRB# IRB00147450) and registered at ClinicalTrials.gov (NCT# 05157074).

### Study Design

This was an open label pilot study. Participants attended the drum-based music therapy intervention group, *The Armstrong Rhythm Method* ^***SM***^ *(ARM)*, a protocol designed and facilitated by Jason Armstrong Baker, a board-certified music therapist with >10 years of clinical experience in rhythm-based intervention techniques. Participants were assigned, based on diagnosis, into two parallel intervention groups: 1) patients diagnosed with PD and their study partners, 2) patients diagnosed with HD and their study partners. Both groups had twice weekly sessions lasting 60 minutes for 12 weeks (a total of 24 sessions). Participants completed assessments at baseline, 6 weeks, and 12 weeks. At the 18-week visit (6 weeks post-intervention completion), participants were asked to complete a Likert-style questionnaire and provide comments regarding their experience with the lessons (the “exit interview”) and to complete the same assessments as at prior visits.

### Study Protocols

All 24 *ARM*^***SM***^ sessions were conducted in a circle to ensure maximum line of sight for each participant. A variety of diverse percussion instruments ranging from hand drums, stick drums, shakers and rhythm sticks were available, as well as adaptive measures to address limited physical functioning. Instrument choice was determined by ease of use for the level of physical ability, skill development needs, and personal participant choice. Participants were informed that they are free to use the same instrument for the duration of each session or to switch instruments at any time.

Utilizing the five components of *ARM*, this protocol guided participants through two stages of engagement to achieve rhythmic skill development and group cohesion for their therapeutic benefit.

#### The Five Components

The five components of the drumming protocol were utilized and arranged into various sequences as determined by the overall needs of the group and the developmental goals.

a. *Call and response rhythms (∼10 minutes):* During this section participants learned rhythms through a simple A-A sequence. This component aimed to improve listening, impulse control (not playing while the leader calls/ waiting to respond), timing, and unison playing with the group (*Figure* 1.1).
b. *Foundational Rhythms and Entrainment (∼20 minutes)*. Using one of the three basic foundational rhythms, the group played one rhythm together in unison. This component aimed to develop unison ensemble playing, sustained attention and listening, group support, and improved endurance through incremental increases in playing duration (*Figure* 1.2).
c. *Stop-time unison rhythmic patterns (∼10 minutes)*. Participants learned how to follow a vocal cue to either stop together while playing a rhythm, or to direct the ensemble to play one-, two- or three-note unison rhythms. This component aimed to improve rhythmic precision, mental focus, impulse control, and ability to follow vocal cues (*Figure* 1.3).
d. *Improvisation and Entrainment (∼10 minutes)*. Participants were invited to improvise, explore and take chances using unison (focused attention) or complimentary (divided attention) rhythms. This component aimed to encourage group support for individual choice and emotional expression, development of rhythmic differentiation between participants, and development of sustained, focused and divided attention.
e. *Rest (∼10 minutes)*. Participants were given time to rest and relax through breathing exercises and simple stretches.

**Figure 1.1.**
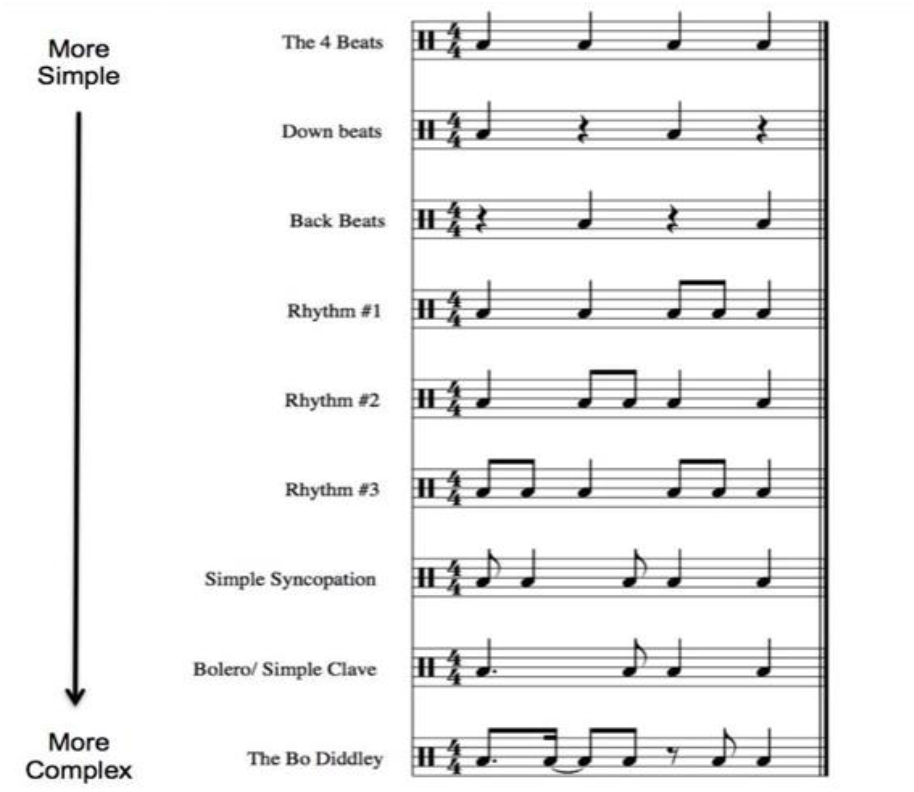
Call and Response Rhythms

**Figure 1.2.**
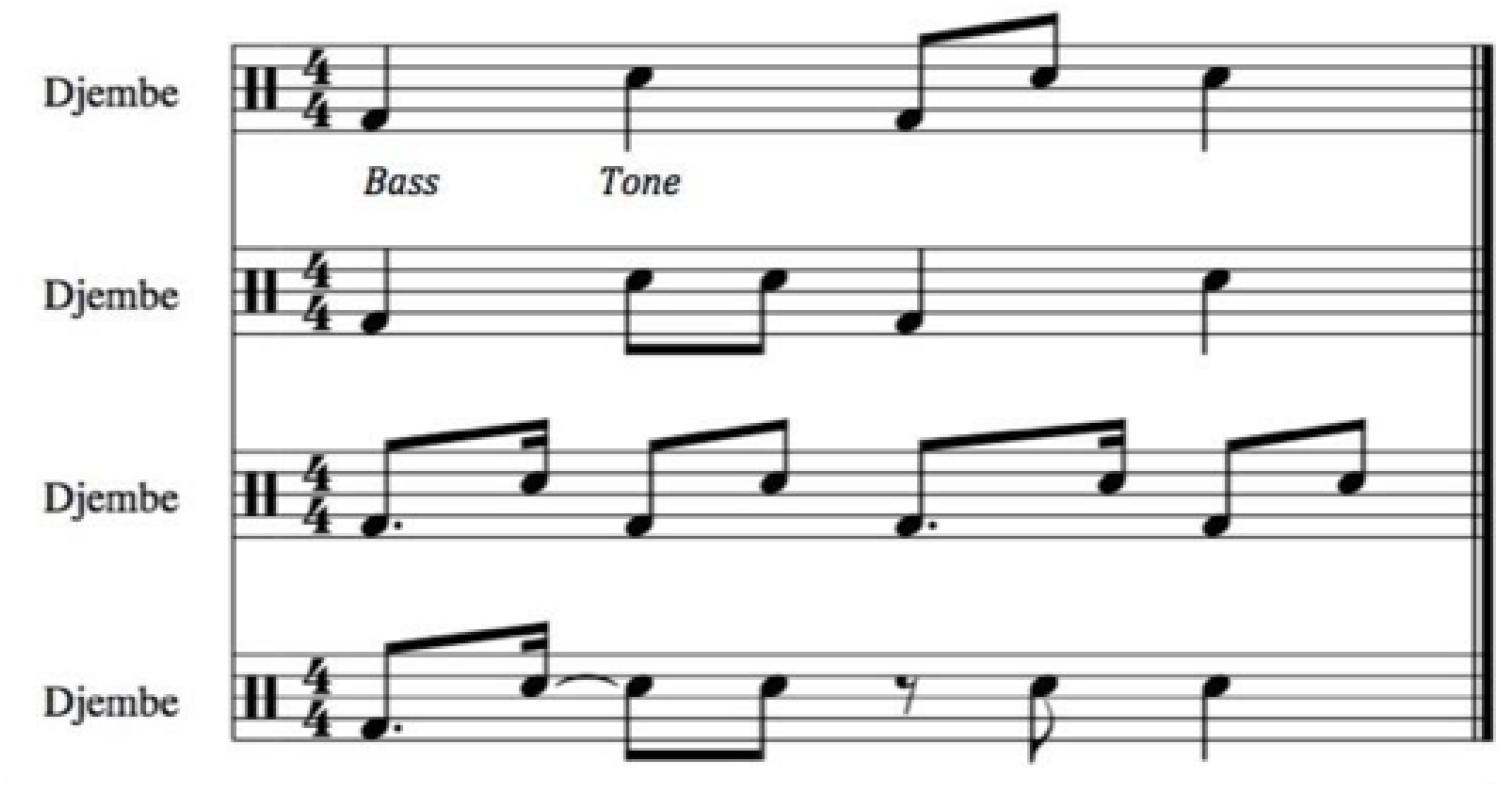
Foundational Rhythms

**Figure 1.3.**
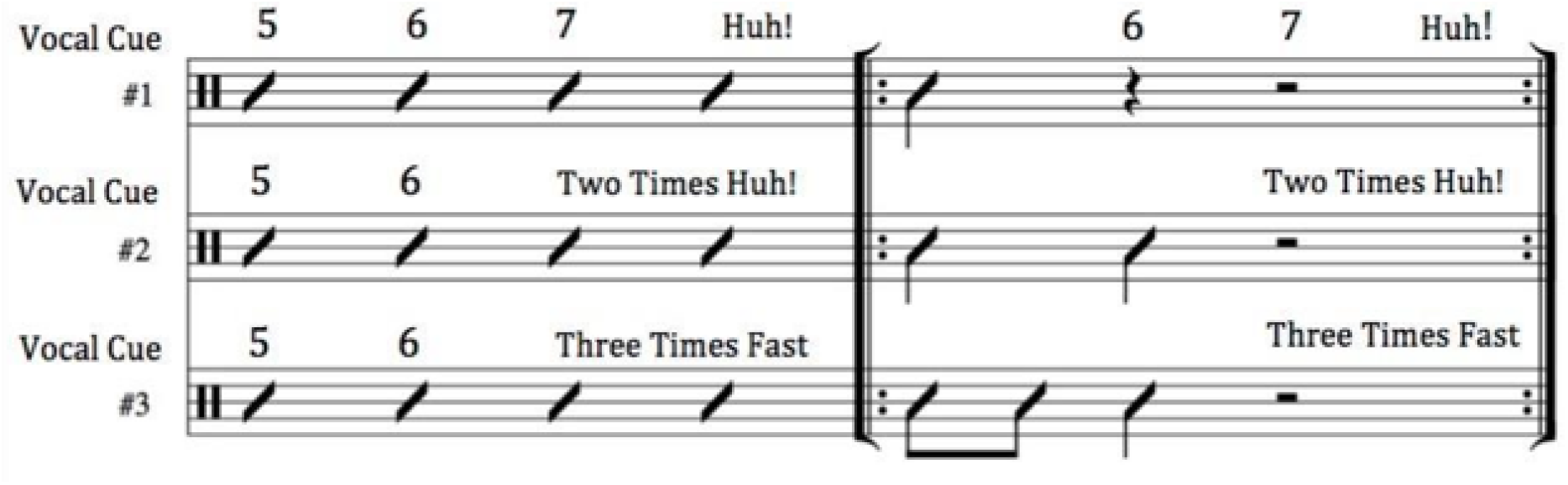
Stop-Time Rhythmic Patterns

**Figure 2.**
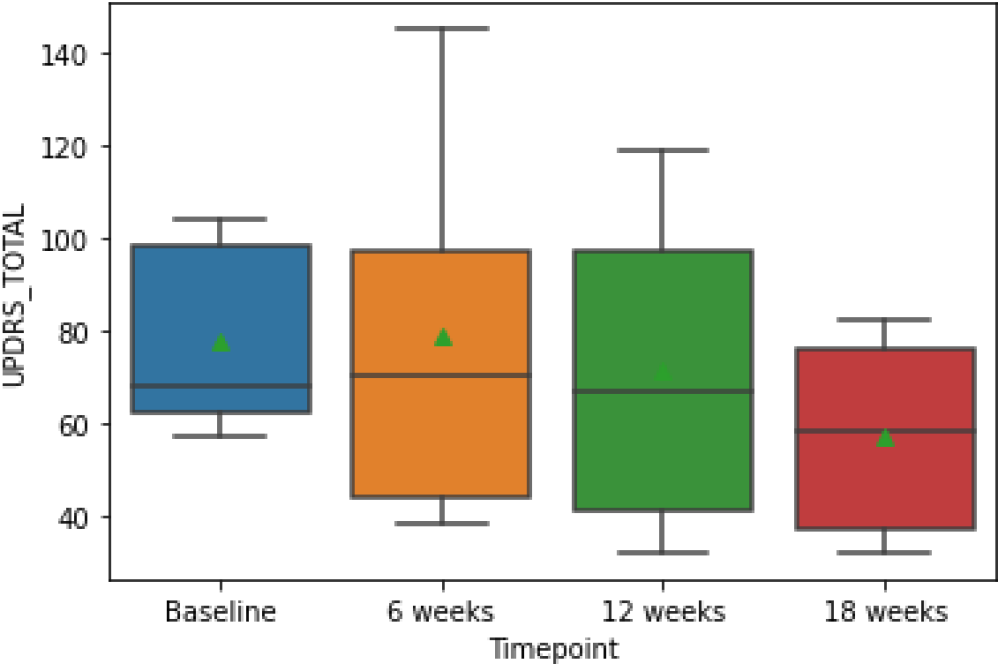
Unified Parkinson’s Disease Rating Scale (UPDRS) Total Scores in Parkinson Disease (PD) participants. Green triangles indicate mean UPDRS total scores. Higher UPDRS total scores indicate greater impairment.

#### The Stages

Over the course of 12 weeks, participants experienced two stages within *ARM*^***SM***^. Both stages of *ARM*^***SM***^ utilized the concept of rhythmic entrainment as a foundational principle to guide participants’ development and growth.

Stage *One:* Introduce the percussion instruments and their safe ergonomic use, present the five components of the protocol, and play simple rhythms.

Stage *Two:* Gradually increase rhythmic complexity, further develop entertainment skills, and improve relative endurance for each participant. More complex rhythms are introduced depending on individual and group ability.

Each phase lasted for one half of the total number of intervention sessions (12 sessions each across a total of 24 sessions). Throughout both phases, instruction was provided to participants in safe hand drumming technique, proper posture, and as needed, appropriate mallets with cushioned handles were provided. Active drum play lasting for periods of 4-7 minutes was balanced with rest opportunities lasting 3-5 minutes (including breathing exercises and stretching) periodically throughout each session. Fatigue was mitigated by gradually increasing the time of sustained physical engagement over the 12-week period by at least 1 minute per week.

### Data Collection and Analysis

The study assessment took place at the same location and during the same calendar week for all participants. MDS-UPDRS (Goetz et al., 2008) and UHDRS (Kieburtz et al., 1996) were used to assess motor symptoms and signs for PD and HD participants, respectively. PD participants completed Parkinson’s Disease Questionnaire (PDQ)-39 (Peto et al., 1995) (scored from 0 to 100; a higher score indicates greater impairment in each dimension) to assess their quality of life. All participants (including caregivers) completed the Neuro-Quality of Life (NeuroQoL) (Cella et al., 2012) at baseline, 6 weeks, 12 weeks, and 18 weeks (6 weeks post-intervention completion). It is a self-report measure of health-related quality of life for people with neurological disorders categorized into 17 subdomains. Our study used the short form in seven subdomains, including social roles and activities, positive affect and well-being, satisfaction with social roles and activities, anxiety, depression, upper limb function, and lower limb function; the raw scores were transformed into T-scores. It is important to note that high scores indicate worse outcomes in anxiety and depression subdomains, while high scores indicate better outcomes in the rest of the subdomains used in this study. Additionally, Short Form 36 (Ware & Sherbourne, 1992) and the Beck Depression Inventory-II (BDI-II) (Beck et al., 1996) were collected for all participants to assess quality of life and depression level, respectively. Zarit Burden Interviews (Zarit et al., 1986) were collected from PD and HD caregivers at baseline and follow up visits. Patients and caregivers also completed a final exit report at 18-week visit (6 weeks post-intervention completion).

### Statistical Analysis

Shapiro-Wilk tests were performed to test for normal distributions among all data groups. Due to non-normality associated with the small sample size, Wilcoxon signed-rank tests were performed to compare the quality-of-life and motor changes between baseline and 6 weeks, baseline and 12 weeks, baseline and 18 weeks, and 12 weeks and 18 weeks. Due to incomplete data collection from one PD patient, 5 of 6 patients’ data were used for this analysis per protocol. A Mann-Whitney U test was conducted to investigate differences between caregivers and patients’ quality-of-life. Four of 6 PD patient-caregiver dyads were included in this analysis due to missing data. Per protocol, one of five dyads’ data were not included as the study partner did not complete the 18-week (follow up) assessments.

### Statistical analysis was performed in Python (version 3.9)

As a result of the smaller size of the Huntington disease group, only descriptive data are reported in the main manuscript. Statistical significance was set to *p* < .05 for all analyses (two-tailed tests). Corrections for multiple comparisons were not performed as this was a pilot study intended to estimate effect sizes.

## Results

Table 1 shows the demographic information for the participants. Participants who completed all assessments at all time points were included in this table.

**Table 1.** Demographic Information.

|  |  | PD | PD Caregivers | HD | HD Caregivers |
| --- | --- | --- | --- | --- | --- |
| Age (mean, (SD)) |  | 73.83 (7.65) | 61.50 (12.99) | 58.33 (11.24) | 53.00 (8.49) |
| Sex | Male | 5 | 0 | 1 |  |
|  | Female | 0 | 5 | 2 | 2 |
| MoCA |  | 25.17 (4.03) | N/A | 23.00 (3.61) | N/A |

For the change scores between PD patients and PD caregivers, there were trend level differences in NeuroQoL outcome (‘Depression’) between PD participants and PD patients (*p* = .05) where PD participants showed increased change of scores (*M*= 5.25, *SD* = 5.72) while PD caregivers indicated decreased change scores (*M* = -3.83, *SD* = 4.48) from baseline to 18 weeks (Figure 3).

**Figure 3.**
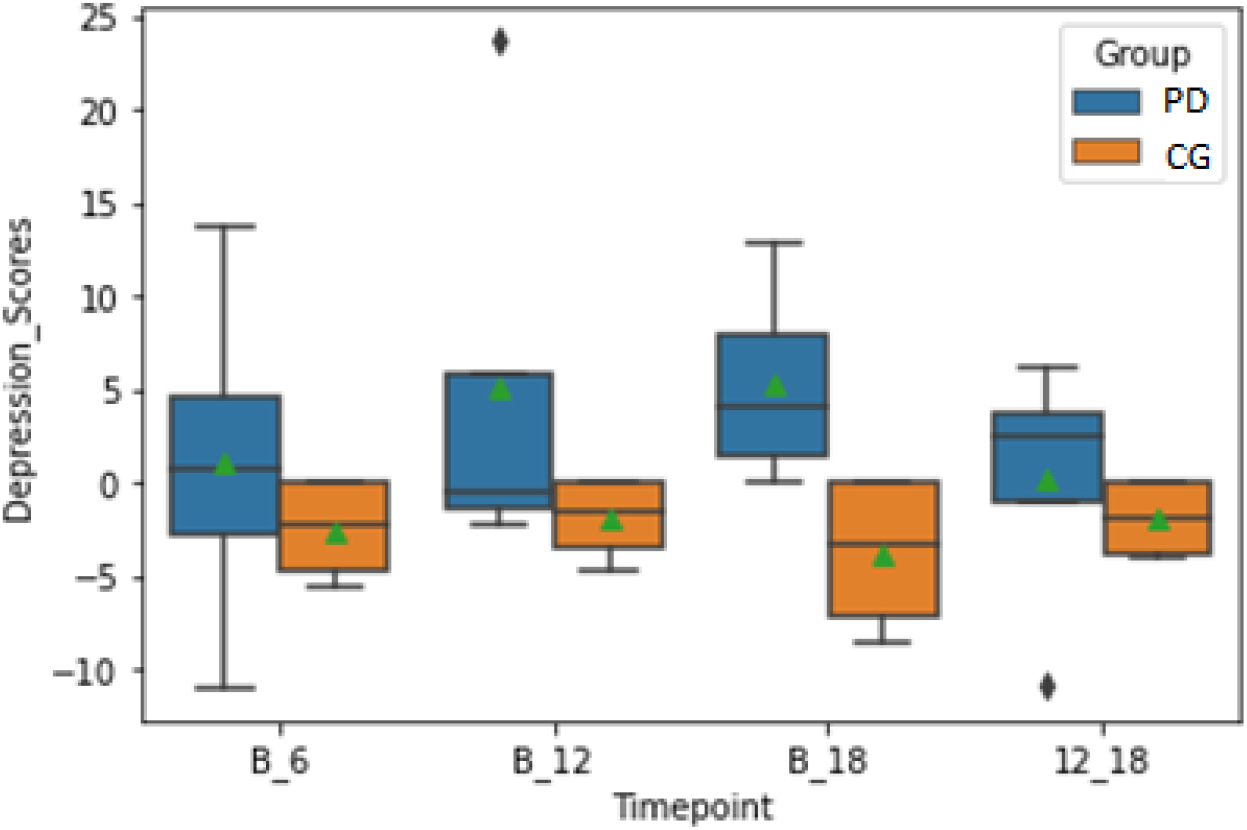
NeuroQoL change scores (Δ) in depression between PD patients and PD caregiver participants. Green triangles indicate mean difference scores of Neuro-QoL. PD = PD patients; CG= Caregivers. B_6= difference between Baseline and 6 weeks (6 weeks – baseline); B_12= difference between Baseline and 12 weeks (12 weeks – baseline); B_18 = difference between Baseline and 18 weeks (18 weeks – baseline); 12_18 = difference between 12 and 18 weeks (18 weeks – 12 weeks). Higher Neurological Quality of Life (NeuroQoL) depression scores indicate more severe depressive symptoms.

Additionally, there was a significant change of social satisfaction scores between PD participants and their caregivers from baseline to 6 weeks, where PD participants showed decreased change of scores (*M* = -2.30, *SD* = 1.64) while PD caregivers showed increased change of scores (*M* = 3.80, *SD* = 3.08), *p* = .02. On the other hand, from 12 weeks to 18 weeks post-baseline, PD participants demonstrated increased change of scores (*M* = 2.53, *SD* = 1.29) while PD caregivers showed decreased change of scores (*M* = -2.10, *SD* = 3.35), *p* = .03 (Figure 4).

**Figure 4.**
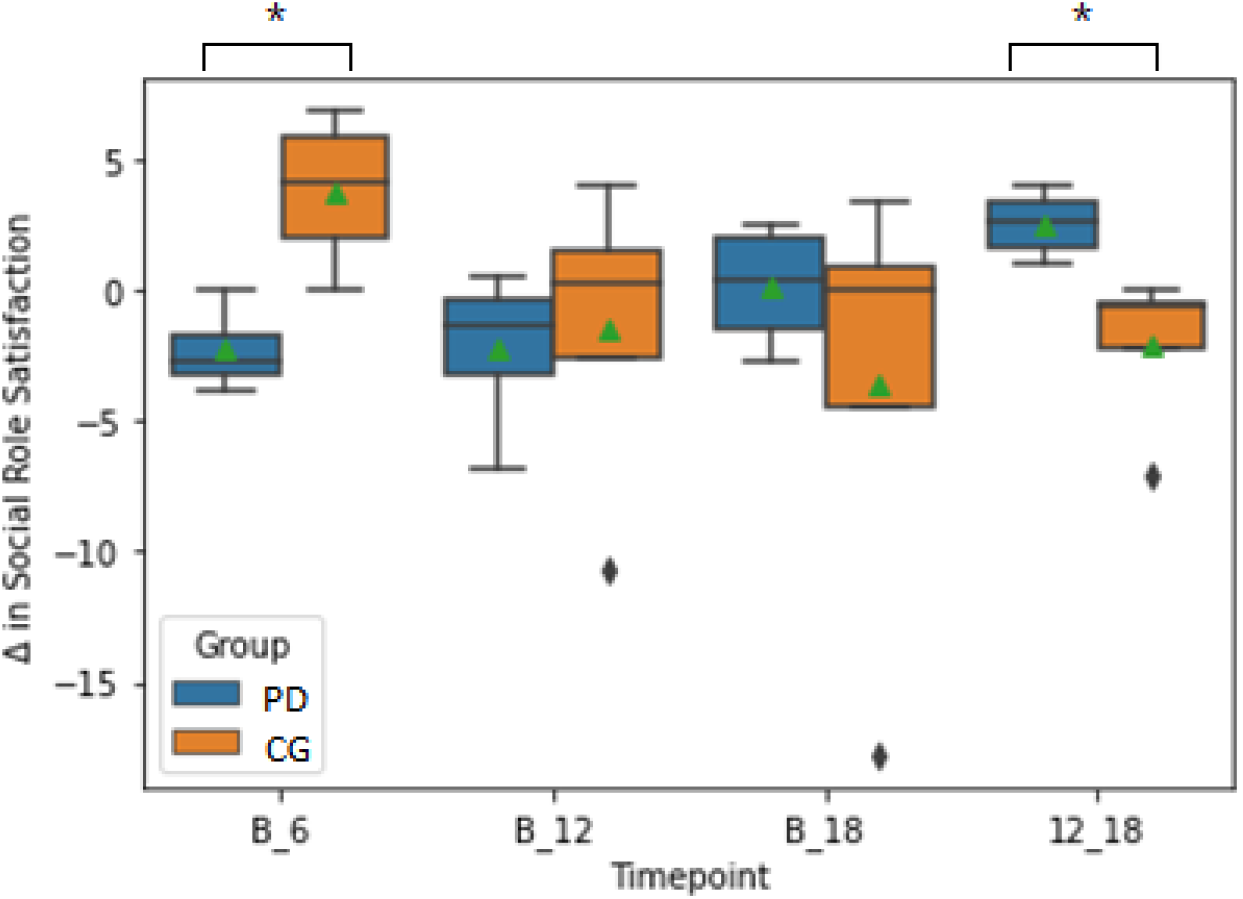
NeuroQoL change scores (Δ) in Social Role Satisfaction between PD patients and PD caregiver Participants. Green triangles indicate mean difference scores of Neuro-QoL in Social Role Satisfaction. PD = PD patients; CG = Caregivers. B_6= difference between Baseline and 6 weeks (6 weeks – baseline); B_12= difference between Baseline and 12 weeks (12 weeks – baseline); B_18 = difference between Baseline and 18 weeks (18 weeks – baseline); 12_18 = difference between 12 and 18 weeks (18 weeks – 12 weeks).

There were no significant differences in PDQ-39 scores for PD patients between baseline (*M* = 40.20, *SD* = 26.29) and 6 weeks (*M* = 43.20, *SD* = 32.31), *p* = 0.48; baseline and 12 weeks (*M* = 36.20, *SD* = 21.09), *p* = .43; and baseline and 18 weeks (*M* = 39.00, *SD* = 28.13), *p* = .76. HD patients and caregiver participants’ descriptive data for NeuroQoL are shown in Table 2.

PD caregivers and HD caregivers’ burden scores did not show any significant changes across timepoints.

Exit questionnaires were collected from a total of 11 participants (PD patients= 4, PD caregivers = 3; HD patients = 2; HD caregivers = 2). All participants who completed this study reported that they very much enjoyed playing drums (4 = very much, 100%) and the social atmosphere (4= very much, 91%). They also described that the class frequency was appropriate (yes = 100%) and that they would attend similar classes (4 = very much, 82%) and recommend them to friends (4 = very much, 91%) in the future.

## Discussion

In this study, after 12 weeks of twice-weekly drum-based music therapy sessions, participants with PD showed noteworthy improvements in several markers of quality of life. It is notable that the previous pilot study (Pantelyat et al., 2016) and the current study show some similar trends, including improvements in motor function and in quality of life. However, the time points at which these trends were observed varied. For motor function, the previous study revealed an initial improvement in motor function evidenced by a decrease in UPDRS scores between baseline and 6 weeks; however, these scores increased between baseline and 12 weeks, indicating a worsening in motor function after drumming interventions were stopped.

The improved UPDRS outcomes in the current study appeared to be maintained throughout the duration of the study, including the final time point 6 weeks after completion of drumming interventions. While the factors impacting this continued improvement were not explored in this pilot study, they merit further attention. One possible explanation for the continued improvement involves a change in the interactions between patients and caregivers.

The interaction of caregivers and patients in the context of therapeutic music interventions for PD and other movement disorders has been examined in several studies previously. Results have shown that music-cued gait training can improve gait stability in patients with Progressive Supranuclear Palsy when administered by a caregiver (Wittwer et al., 2019). Patients with dementia showed decreased disruptiveness (Ridder et al., 2013) as well as decreased resistiveness to care and increased positive emotions when administered music-based interventions by a caregiver (Hammar et al., 2011). Additionally, caregivers of people with PD have reported benefit from joint participation in activities such as music therapy (Prado et al., 2020). Group therapy sessions have been shown to benefit people with PD. For example, several studies have shown that singing in a group setting can improve speaking quality for participants with PD (Elefant et al., 2012; Yinger & Lapointe, 2012). In light of previous studies showing the importance of caregiver involvement as well as others demonstrating that instrument playing enhances social cohesion (Waldon, 2001) and reduces participant-reported burden and anxiety levels (Hilliard, 2001), our findings provide insight into the potency of combining instrument playing, group music therapy sessions, and caregiver participation. In order to further probe the importance of combining these elements in treating PD, future studies should continue to investigate the effects of providing PD patients with a combination of instrument-based, group music therapy sessions that include their caregivers.

In our study, it appeared that both motor function and social satisfaction were maintained by patients with PD for longer time period than a previous pilot study (Pantelyat et al., 2016), suggesting that 12 weeks of drumming may provide additional benefits vs 6 weeks of drumming. Future music therapy studies should further investigate the role of caregivers in facilitating long-lasting improvements in PD patients by having caregivers participate in interventions alongside patients.

It is important to note that while the participants showed improved overall motor function (as measured by UPDRS total scores) as well as increased social satisfaction, PD caregivers reported *decreased* social satisfaction scores between baseline and 6 weeks, possibly because participation in drumming took them away from other preferred activities. Interestingly, we observed a dissociation of score changes between completion of drumming and (12 weeks) and the final follow up (18 weeks): PD participants reported decreased social satisfaction and caregivers reported increased social satisfaction between 12 weeks and 18 weeks. This may have occurred because PD patients were missing the social interactions of group drumming, while their caregivers appreciated getting back additional time (2 hours per week, excluding travel) for their preferred activities after drumming was completed. In turn, this points to *a priori* differences in motivation/reasons to enroll in our study between patients and caregivers. Further exploration of the dissociated pattern of outcomes between PD patients and their caregivers noted in our group drumming music therapy intervention is important, in addition to investigating changes in quality of life. By gaining a better understanding of these distinct perceptions, music therapists and others administering music-based interventions may be able to modify their approach towards patients and caregivers to provide tailored support that maximizes positive impact.

In this study, the participants were asked to complete a simple exit interview questionnaire to obtain their insights into the drum sessions (available in Supplementary Materials, Table 4). However, more extensive qualitative written responses to sessions would provide an excellent source of information to develop the program in the future. Furthermore, collecting information regarding participants’ musical backgrounds and prior musical experiences will provide a better understanding of the relationship between prior musical experiences and intervention outcomes.

The key limitation for the current study is its small sample size, especially for the HD group. Due to this, HD patient results were only used for descriptive data, leaving data from the PD patient-caregiver group to be used for data analysis. This small number of participants, combined with the small number of caregivers, severely limits the generalizability of our results. Additionally, this was an open-label study and thus any detected improvements cannot be definitively attributed to the study intervention. In spite of this, considering the paucity of published group intervention studies in PD and HD using music therapy for both patients and their caregivers, our study can serve as a valuable resource for estimating the appropriate sample size in appropriately powered controlled studies. To achieve a power of 0.80 with an alpha level of 0.05, the necessary sample size was determined using Cohen’s d values through power calculations with G*Power, version 3.1.9.7. For MDS-UPDRS total scores, the comparison between baseline and 18 weeks had an effect size of d = 0.94, which indicated that a total of 12 participants would be needed to detect differences at an alpha of 0.05 with a power of 0.80. In the case of changes in social satisfaction scores (NeuroQoL) from baseline to 6 weeks, 4 participants in each group (a total of 8) with a d value of -2.47 were required for comparisons between PD patients and PD caregivers. For the depression domain, 7 participants in each group (a total of 14) with a d value of 1.77 would be needed to compare changes in scores from baseline to 18 weeks between PD patients and PD caregivers (see Supplementary Materials, Table 5).

In summary, this open label pilot parallel group study of drumming was not powered to detect significant changes, and there were not enough HD participants enrolled to perform meaningful statistical analysis. Despite this, we found positive motor and quality of life changes in the PD group. In addition, there was a dissociation between PD patients and caregivers’ reported social satisfaction in the 6 weeks after drumming intervention (weeks 12 to 18), suggesting that caregivers were impacted differently by their participation in the study and may have had different motivations for study participation. Our study warrants further exploration of the impact of group drumming interventions on patients with PD and HD and their care partners.

## Data Availability

All data produced in the present study are available upon reasonable request to the authors

## Declarations

The authors declare that there are no disclosures to report.

## Data Availability Statement

Raw data are available upon request addressed to the corresponding author.

## Funding Statement

No external funding was received for this study.

### Acknowledgement

We thank all study participants for their engagement in our work.

## Author Roles

Note that some submission types have a limit for the number of authors permitted. At the end of the manuscript, list the itemized contributions in number/letter format, as below. These should include but are not restricted to:

1. Research project: A. Conception, B. Organization, C. Execution;
2. Statistical Analysis: A. Design, B. Execution, C. Review and Critique;
3. Manuscript Preparation: A. Writing of the first draft, B. Review and Critique;

LW: 1B, 1C, 2C, 3A, 3B

KK: 2A, 2B, 2C, 3A, 3B

GD: 2B, 2C, 3B

CB: 3A, 3B

KD: 1A, 1B, 3B

DL: 1B, 1C, 3B

GS-C: 1C, 3B

JAB: 1B, 1C, 3B

SB-G: 1B, 3B

JB: 1A, 1B, 2C, 3B

AP: 1A, 1B, 2A, 2C, 3B

