## Supplementary Materials for "DRUM-PD/HD: The Impact of a Pilot Group Drum-Based Music Therapy Intervention on Quality of Life and Motor Symptoms in Parkinson Disease and Huntington Disease"

**Table 1. A comparison of NeuroQoL scores between time points in PD patients and their caregivers**

|  |  | Patients | | | Caregivers | | |
| --- | --- | --- | --- | --- | --- | --- | --- |
|  | Time point | n | Mean | SD | n | Mean | SD |
| NeuroQoL-Anxiety | Baseline | 5 | 44.84 | 5.82 | 5 | 45.44 | 10.70 |
|  | 6 weeks | 5 | 46.76 | 6.18 | 5 | 51.68 | 17.04 |
|  | 12 weeks | 5 | 45.82 | 6.98 | 5 | 44.16 | 7.90 |
|  | 18 weeks | 5 | 47.06 | 3.22 | 4 | 47.40 | 9.33 |
| NeuroQoL-Depression | Baseline | 5 | 42.98 | 5.65 | 5 | 46.06 | 10.22 |
|  | 6 weeks | 5 | 44.36 | 6.98 | 5 | 42.38 | 8.39 |
|  | 12 weeks | 5 | 44.86 | 9.76 | 5 | 42.84 | 8.85 |
|  | 18 weeks | 5 | 46.96 | 2.91 | 4 | 42.43 | 7.51 |
| NeuroQoL-Upper Limb | Baseline | 5 | 41.12 | 12.72 | 5 | 50.46 | 7.47 |
|  | 6 weeks | 5 | 39.16 | 8.93 | 5 | 51.78 | 4.52 |
|  | 12 weeks | 5 | 44.28 | 9.39 | 5 | 51.78 | 4.52 |
|  | 18 weeks | 5 | 41.16 | 12.26 | 4 | 51.28 | 5.05 |
| NeuroQoL-Lower Limb | Baseline | 5 | 43.62 | 13.11 | 5 | 53.64 | 4.65 |
|  | 6 weeks | 5 | 39.28 | 7.58 | 5 | 52.60 | 9.82 |
|  | 12 weeks | 5 | 45.40 | 9.86 | 5 | 53.74 | 7.45 |
|  | 18 weeks | 5 | 45.74 | 13.68 | 4 | 50.90 | 5.69 |
| NeuroQoL-Social Functioning | Baseline | 5 | 45.46 | 9.49 | 5 | 48.3 | 10.97 |
|  | 6 weeks | 5 | 45.20 | 7.53 | 5 | 53.04 | 10.05 |
|  | 12 weeks | 5 | 44.70 | 6.22 | 5 | 47.28 | 9.79 |
|  | 18 weeks | 5 | 45.96 | 9.22 | 4 | 50.73 | 11.30 |
| NeuroQoL-Positive Affect | Baseline | 5 | 55.56 | 9.35 | 5 | 57.32 | 12.63 |
|  | 6 weeks | 5 | 53.90 | 7.70 | 5 | 59.56 | 10.40 |
|  | 12 weeks | 5 | 54.94 | 5.76 | 5 | 61.48 | 10.65 |
|  | 18 weeks | 5 | 54.50 | 11.44 | 4 | 59.58 | 11.65 |
| NeuroQoL-Social Satisfaction | Baseline | 5 | 45.64 | 4.34 | 5 | 48.92 | 8.65 |
|  | 6 weeks | 5 | 43.68 | 3.64 | 5 | 54.40 | 8.95 |
|  | 12 weeks | 5 | 43.56 | 3.59 | 5 | 49.38 | 7.77 |
|  | 18 weeks | 5 | 45.48 | 4.55 | 4 | 47.18 | 9.69 |

Notes: NeuroQoL anxiety and depression: High scores indicate worse (undesirable) self-reported health

**Table 2. NeuroQoL scores between time points in HD patients and their caregivers**

|  |  | Huntington’s Disease (HD) | | | | | |
| --- | --- | --- | --- | --- | --- | --- | --- |
|  |  | Patients | | | Caregivers | | |
|  | Time point | n | Mean | SD | n | Mean | SD |
| NeuroQoL-Anxiety | Baseline | 3 | 50.53 | 12.27 | 2 | 50.45 | 6.44 |
|  | 6 weeks | 3 | 55.23 | 4.44 | 2 | 46.60 | 14.42 |
|  | 12 weeks | 3 | 52.97 | 9.41 | 2 | 47.70 | 7.92 |
|  | 18 weeks | 3 | 64.27 | 10.86 | 2 | 37.40 | 15.56 |
| NeuroQoL-Depression | Baseline | 3 | 48.53 | 10.10 | 2 | 46.45 | 4.74 |
|  | 6 weeks | 3 | 48.83 | 5.09 | 2 | 44.85 | 11.24 |
|  | 12 weeks | 3 | 48.80 | 10.37 | 2 | 44.10 | 10.18 |
|  | 18 weeks | 3 | 47.27 | 8.98 | 2 | 49.55 | 17.89 |
| NeuroQoL-Upper Limb | Baseline | 3 | 47.07 | 5.83 | 2 | 58.60 | 0 |
|  | 6 weeks | 3 | 40.30 | 11.73 | 2 | 58.60 | 0 |
|  | 12 weeks | 3 | 42.53 | 10.66 | 2 | 58.60 | 0 |
|  | 18 weeks | 3 | 40.60 | 11.84 | 2 | 58.60 | 0 |
| NeuroQoL-Lower Limb | Baseline | 3 | 47.03 | 10.03 | 2 | 58.60 | 0 |
|  | 6 weeks | 3 | 45.67 | 11.48 | 2 | 58.60 | 0 |
|  | 12 weeks | 3 | 47.90 | 9.64 | 2 | 58.60 | 0 |
|  | 18 weeks | 3 | 46.73 | 10.28 | 2 | 58.60 | 0 |
| NeuroQoL-Social Functioning | Baseline | 3 | 46.93 | 5.69 | 2 | 60.20 | 0 |
|  | 6 weeks | 3 | 47.10 | 12.08 | 2 | 48.85 | 1.91 |
|  | 12 weeks | 3 | 47.33 | 11.77 | 2 | 53.85 | 8.98 |
|  | 18 weeks | 3 | 46.90 | 11.52 | 2 | 55.90 | 6.08 |
| NeuroQoL-Positive Affect | Baseline | 3 | 51.87 | 4.47 | 2 | 57.00 | 8.91 |
|  | 6 weeks | 3 | 50.40 | 9.70 | 2 | 58.95 | 12.80 |
|  | 12 weeks | 3 | 53.53 | 12.53 | 2 | 56.20 | 16.69 |
|  | 18 weeks | 3 | 42.87 | 6.15 | 2 | 53.85 | 13.36 |
| NeuroQoL-Social Satisfaction | Baseline | 3 | 45.13 | 6.27 | 2 | 52.00 | 0 |
|  | 6 weeks | 3 | 49.47 | 9.64 | 2 | 50.90 | 1.56 |
|  | 12 weeks | 3 | 47.30 | 11.44 | 2 | 50.90 | 1.56 |
|  | 18 weeks | 3 | 48.97 | 10.00 | 2 | 47.00 | 3.96 |

**Table 3. A comparison of NeuroQoL scores changes between PD patients and their caregivers**

|  |  | Patients | | Caregivers | |  |
| --- | --- | --- | --- | --- | --- | --- |
|  | Changes (Δ) | Mean | SD | Mean | SD | *p*-value |
| NeuroQoL-Anxiety | Baseline-6 weeks | 1.43 | 12.17 | 0.08 | 2.82 | 0.47 |
|  | Baseline-12 weeks | 2.80 | 10.31 | -2.50 | 4.00 | 0.66 |
|  | Baseline-18 weeks | 2.25 | 6.65 | 2.08 | 14.33 | 0.88 |
|  | 12 weeks -18 weeks | -0.55 | 4.81 | 4.58 | 11.75 | 0.66 |
| NeuroQoL-Depression | Baseline-6 weeks | 1.05 | 10.11 | -2.50 | 2.92 | 0.46 |
|  | Baseline-12 weeks | 5.10 | 12.43 | -1.93 | 2.33 | 0.46 |
|  | Baseline-18 weeks | 5.25 | 5.72 | -3.82 | 4.48 | 0.05 |
|  | 12 weeks -18 weeks | 0.15 | 7.50 | -1.9 | 2.12 | 0.30 |
| NeuroQoL-Upper Limb | Baseline-6 weeks | -3.55 | 11.53 | 1.65 | 3.30 | 0.62 |
|  | Baseline-12 weeks | 1.88 | 2.39 | 1.65 | 3.30 | 0.87 |
|  | Baseline-18 weeks | -0.85 | 0.99 | 1.65 | 3.30 | 0.14 |
|  | 12 weeks -18 weeks | -2.73 | 3.26 | 0 | 0 | 0.19 |
| NeuroQoL-Lower Limb | Baseline-6 weeks | -5.65 | 6.79 | -1.3 | 8.30 | 0.47 |
|  | Baseline-12 weeks | 2.35 | 2.91 | -1.73 | 3.45 | 0.14 |
|  | Baseline-18 weeks | 2.88 | 4.80 | -3.35 | 4.71 | 0.09 |
|  | 12 weeks -18 weeks | 1.38 | 6.13 | -1.62 | 5.82 | 0.88 |
| NeuroQoL-Social Functioning | Baseline-6 weeks | -0.15 | 5.25 | 1.55 | 2.65 | 0.77 |
|  | Baseline-12 weeks | 0.23 | 3.44 | -2.30 | 7.98 | 0.69 |
|  | Baseline-18 weeks | 3.13 | 5.22 | 1.03 | 2.54 | 0.66 |
|  | 12 weeks -18 weeks | 2.10 | 5.60 | 3.33 | 6.07 | 0.89 |
| NeuroQoL-Positive Affect | Baseline-6 weeks | -5.15 | 8.21 | 2.80 | 3.46 | 0.19 |
|  | Baseline-12 weeks | -3.30 | 6.17 | 3.53 | 4.43 | 0.18 |
|  | Baseline-18 weeks | 2.70 | 7.73 | 3.25 | 3.97 | 0.66 |
|  | 12 weeks -18 weeks | 3.68 | 8.42 | -0.28 | 0.55 | 0.30 |
| NeuroQoL_Social Satisfaction | Baseline-6 weeks | -2.30 | 1.64 | 3.78 | 3.05 | 0.04* |
|  | Baseline-12 weeks | -2.28 | 3.20 | -1.53 | 6.37 | 0.49 |
|  | Baseline-18 weeks | 0.13 | 2.51 | -3.63 | 9.58 | 1 |
|  | 12 weeks -18 weeks | 2.53 | 1.34 | -2.10 | 3.35 | 0.03* |

*Note*: **p*<.05

**Table 4. Exit Questionnaire**

| **Questions** |  |
| --- | --- |
| Enjoyed playing drums | 0 1 2 3 4 |
| Able to follow instructions | 0 1 2 3 4 |
| Enjoyed social atmosphere | 0 1 2 3 4 |
| Convenient to attend | 0 1 2 3 4 |
| Class helped arms/hand move | 0 1 2 3 4 |
| Would recommend class to friends | 0 1 2 3 4 |
| Would attend similar class in future | 0 1 2 3 4 |
| Class frequency is appropriate | 0 1 2 3 4 |
| Plans to seek music instruction as rec activity | 0 1 2 3 4 |
| Notify for similar research projects | 0 1 2 3 4 |

**Table 5. Sample Size Estimation**

| **Assessments** | **Domain** | **Comparison** | **Effect size**  **(Cohen’s *d*)** | **Estimated**  **Sample size** |
| --- | --- | --- | --- | --- |
| NeuroQoL | Social Satisfaction | PD Participants vs PD caregivers (Δbaseline-6 weeks): Mann-Whitney | -2.47 | 8 (4 per group) |
|  |  | PD Participants vs PD caregivers (Δ12-18 weeks): Mann-Whitney | 1.82 | 14 (7 per group) |
|  | Depression | PD Participants vs PD caregivers (Δbaseline-18 weeks): Mann-Whitney | 1.77 | 14 (7 per group) |
| UPDRS | Total | PD patients’ baseline vs PD patients’ 18 weeks: Wilcoxon’s | 0.94 | 12 |

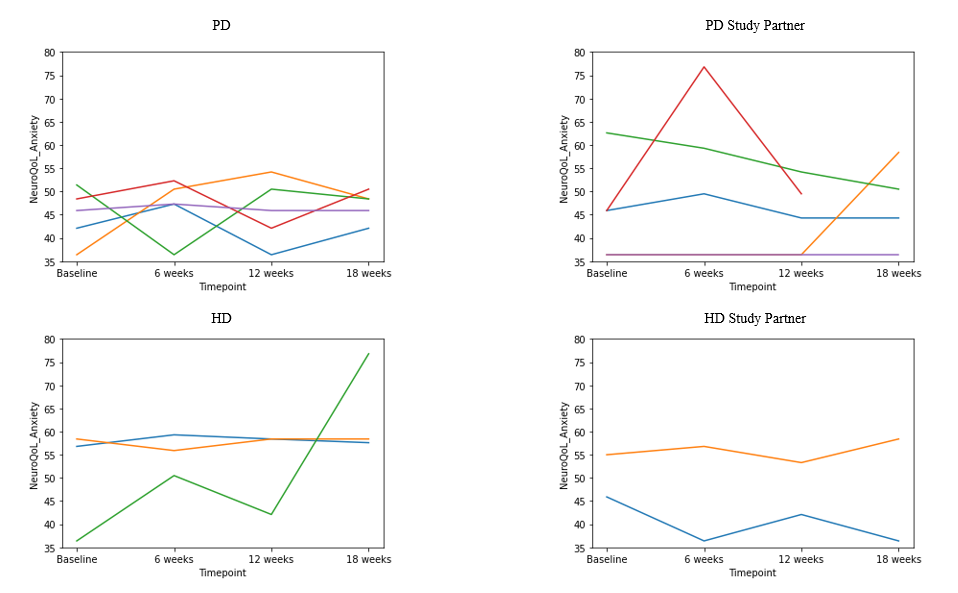

*Figure 1*. Individual Spaghetti plots for NeuroQoL (Anxiety).

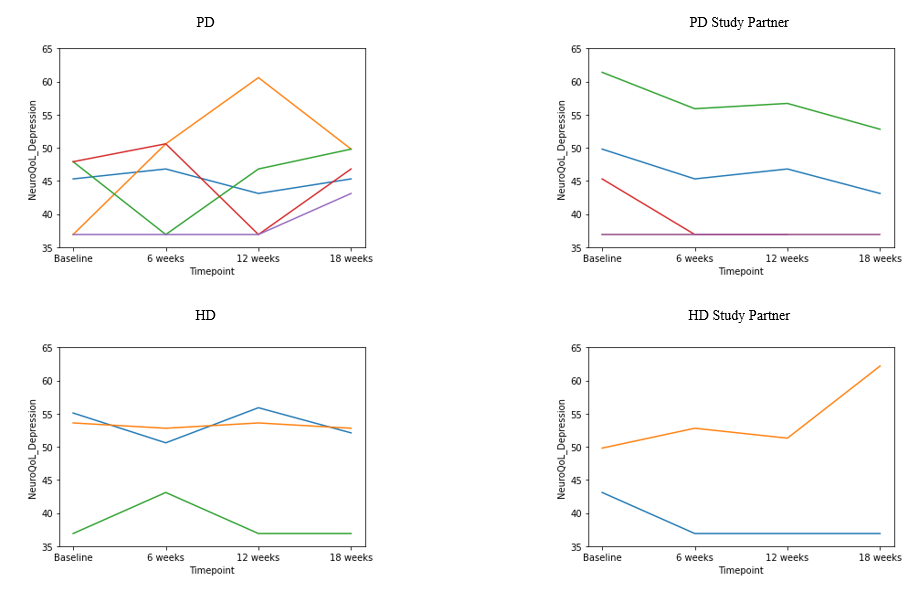

*Figure 2.* Individual Spaghetti plots for NeuroQoL (Depression).

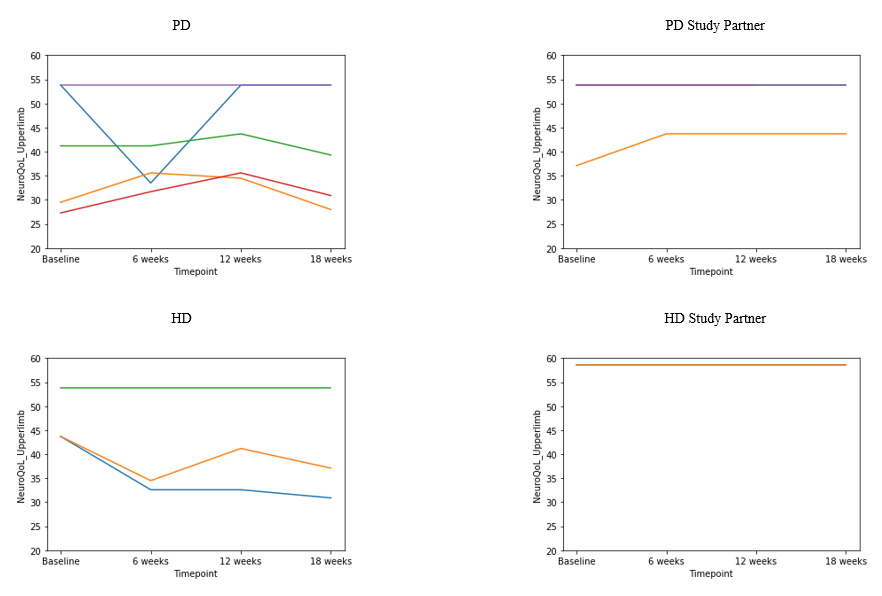

*Figure 3*. Individual Spaghetti plots for NeuroQoL (Upper limb).

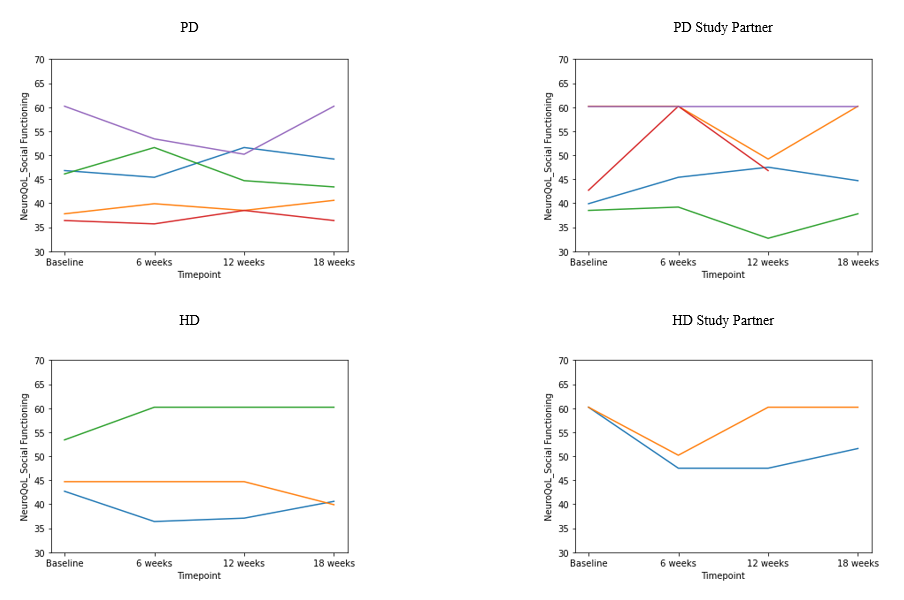

*Figure 4*. Individual Spaghetti plots for NeuroQoL (Social Functioning).

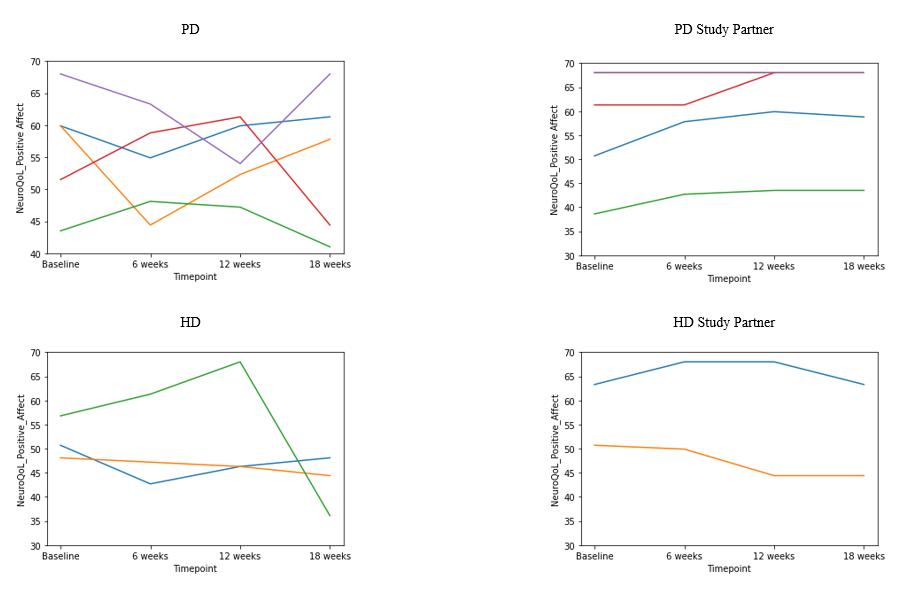

*Figure 5*. Individual Spaghetti plots for NeuroQoL (Positive Affect).

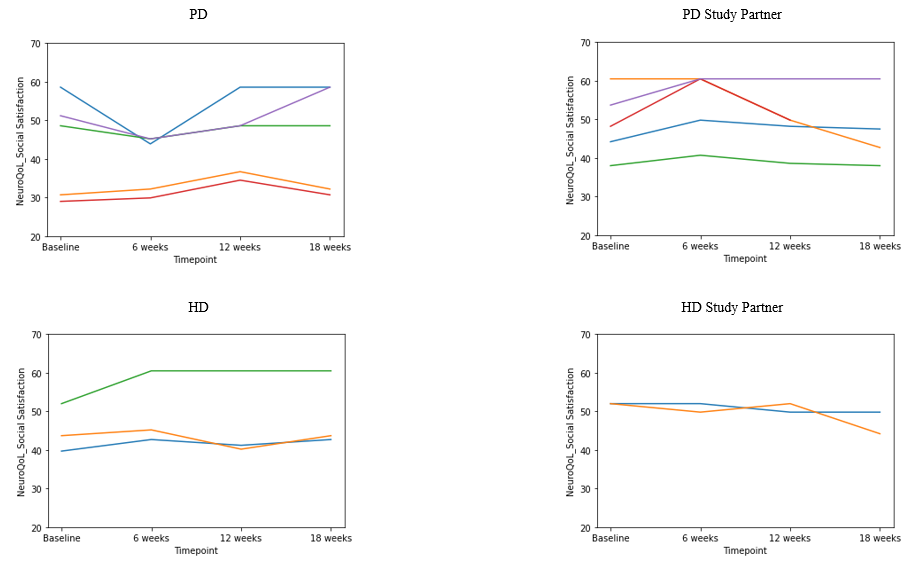

*Figure 6.* Individual Spaghetti plots for NeuroQoL (Social Role Satisfaction).
